# COVID-19 is rapidly changing: Examining public perceptions and behaviors in response to this evolving pandemic

**DOI:** 10.1101/2020.05.04.20091298

**Authors:** H Seale, AE Heywood, J Leask, M Sheel, S Thomas, D.N Durrheim, K Bolsewicz, R Kaur

**Affiliations:** School of Public Health and Community Medicine, University of New South Wales, Sydney, NSW, Australia; Faculty of Medicine and Health, University of Sydney, Sydney, NSW, Australia; National Centre for Immunisation Research and Surveillance, Kids Research, Sydney Children’s Hospitals Network, Westmead, NSW, Australia; National Centre for Epidemiology and Population Health, Research School of Population Health, ANU College of Health and Medicine, The Australian National University, Acton, ACT, Australia; School of Medicine and Public Health, University of Newcastle, Wallsend, NSW, Australia; Office of Medical Education, Faculty of Medicine, University of New South Wales, Sydney, NSW, Australia

## Abstract

**Background:** Since the emergence of SARS-CoV-2, the virus that causes coronavirus disease (COVID-19) in late 2019, communities have been required to rapidly adopt community mitigation strategies rarely used before, or only in limited settings. This study aimed to examine the attitudes and beliefs of Australian adults towards the COVID-19 pandemic, and willingness and capacity to engage with these mitigation measures. In addition, we aimed to explore the psychosocial and demographic factors that are associated with adoption of recommended hygiene-related and avoidance-related behaviors.

**Methods:** A national cross-sectional online survey of 1420 Australian adults (18 years and older) was undertaken between the 18 and 24 March 2020. The statistical analysis of the data included univariate and multivariate logistic regression analysis.

**Findings:** The survey of 1420 respondents found 50% (710) of respondents felt COVID-19 would ‘somewhat’ affect their health if infected and 19% perceived their level of risk as high or very high. 84·9% had performed ≥1 of the three recommended hygiene-related behaviors and 93·4% performed ≥1 of six avoidance-related behaviors over the last one month. Adopting avoidance behaviors was associated with trust in government/authorities (aOR: 5·5, 95% CI 3-9·0), higher perceived rating of effectiveness of behaviors (aOR: 4·3, 95% CI: 2·8-6·9) and higher levels of perceived ability to adopt social distancing strategies (aOR: 1·8, 95% CI 1·1-3·0).

**Interpretation:** In the last two months, members of the public have been inundated with messages about hygiene and social (physical) distancing. However, our results indicate that a continued focus on supporting community understanding of the rationale for these strategies, as well as instilling community confidence in their ability to adopt or sustain the recommendations is needed.

**Funding:** None

## INTRODUCTION

In the course of four months, since the first reports about a novel strain of coronavirus, severe acute respiratory syndrome coronavirus 2 (SARS-CoV-2) emerging in December 2019, countries around the world have introduced a range of community mitigation strategies with the aim to lower the trajectory of this pandemic by reducing transmission, and avoid overwhelming health services. Community mitigation strategies refer to measures that people, and communities can take to slow the spread of infection during a period when vaccines and/or medical treatments that are not available [1]. They include the use of personal protective measures for everyday use (e.g., voluntary home isolation of ill persons, respiratory etiquette, and hand hygiene); community measures aimed at increasing social distancing (e.g., maintaining a physical distance of 1·5-2·0 meters between people, staying at home and postponing or cancelling gatherings); and environmental measures (e.g., routinely disinfecting surfaces). In some settings these strategies are voluntary, whereas in others they are now enforced (such as, via fines and/or jail time).

Governments are implementing strategies at large-scale that have previously been used in limited ways and for limited time periods i.e. during Ebola, avian influenza outbreaks and SARS. This means a large proportion of the population do not have prior experience undertaking these strategies. People’s ability to comply with recommendations during emergency situations is influenced by a range of modifiable and nonmodifiable factors including: (1) what people perceive their susceptibility to infection to be[2]; (2) whether they perceive the infection to be serious, if acquired; (3) whether they have the necessary capacity, confidence and resources to comply with the strategies[3]; and (4) their sociodemographic status [4]. Effective control of this pandemic requires an understanding of people’s perceptions about their willingness, motivation and ability/capacity to adopt strategies and how this relates to their perceived risk [5]. Perceived costs, perceptions about the benefits of the behaviors and the perceived impact of an individual’s behavior on another’s health will also influence engagement with these behaviors [6, 7].

In Australia, the government has recommended specific behaviors that can be classed as hygiene-related and avoidance related. To engage in these behaviors, people will weigh up the perceived costs and benefits related to themselves and others. It is therefore important to understand community perceptions and behaviors in order to develop effective messages. Accordingly, we carried out a cross sectional online survey of a large, demographically representative sample of the population of Australia in March 2020.

## METHODS

### Cross sectional online survey

We conducted an online survey of Australian residents via a market research company (Quality Online Research (QOR)) between 18 and 24 March 2020. This sample size provided us with a sample error of ±3%. Proportional quota sampling was used to ensure that respondents were demographically representative of the general public, with quotas based on age, gender and state/territory. Respondents were required to be 18 years or older and to speak English. Respondents earned points for completing the survey. After reading the participant information, consent was implied if the person completed the survey and submitted it via the QOR website. Ethics approval for the study was obtained from the University of New South Wales (HC200190).

### Survey design

The questions for this survey were adapted from published studies by HS during the 2009 influenza H1N1/A pandemic [8, 9]. The study tool is available upon request. Ten items were used to assess respondent perceptions about the COVID-19 pandemic, including perceived risk level and impact on health (if infected). 8/10 items were phrased as statements, with Likert response options scored as 5 for strongly agree through to 1 for strongly disagree. Two items measuring participants’ level of worry about current Covid-19 were used on a 5 point Likert scale ranging from 1 for strongly disagree to 5 for strongly agree, these were combined and changed into a dichotomous scale of high and low.

Respondents were asked to rate the perceived level of effectiveness of 13 items in reducing the risk from COVID-19 on a 5-point scale These items included those promoted by the government and those that were not (mask use when not symptomatic, taking antibiotics). The strategies were grouped into: (1) hygiene related behaviors (hand washing/sanitizing, cleaning surfaces) and (2) avoidance-related behaviors (avoiding crowds, public transport, and complying with quarantine restrictions). Given the relative novelty of social distancing for the Australian community, we also included a question that assessed the respondent’s ability to adopt 6 different social distancing strategies (working from home, keeping children home from school, avoiding travelling, avoiding large crowds, quarantine if exposed, and isolation if symptomatic) with possible response options scored as 1 for very high and 5 for very low. The last section of the survey included six items focused on self-isolation. Respondents were asked to comment on their willingness to comply, their level of concern regarding the impact on being placed into self-isolation (at home), their ability to comply, their access to assistance from family/friends and issues they have with the strategy. All predictor variables and the items and scales are described in the supplementary materials.

We collected data on gender, age, education and employment status, children (including attendance at childcare/school), country of birth/language spoken at home, whether they identify as Aboriginal and/or Torres Strait Islander, international travel patterns since 1 January 2020, private healthcare insurance coverage, income protection insurance, the presence of any chronic illness and self-reported health status (very good, good, moderate, poor, very poor). Due to the promotion of social distancing and working from home by the Australian Government, we also included two items that assessed access to internet and a computer at home.

### Analysis

Two primary outcome variables used were hygiene-related and avoidance-related behaviors. (see Table 1). Cronbach’s alpha was calculated to check the internal consistency of the combined items within predictor variable measured using same scale (See supplementary materials). Univariate associations were tested between primary outcome measures and demographic factors. Univariate associations between worry and outcome measures was also assessed. Two separate multivariate logistic regression models were used to measure the associations of perception factors with each outcome factor after controlling for demographics and worry variables. Demographic variables with a P<0.25 in the univariate analysis were used to adjust the models.

**Table 1:** Adoption of hygiene-related and avoidance-related behaviors.in response to COVID-19.

|  | Number (%) |
| --- | --- |
| **Hygiene-related behaviors: actions taken over last month due to COVID-19 (Cronbach's alpha=0.701)** |  |
| Increased the time I spent cleaning or disinfecting things I might touch, such as door knobs | 537/1420 (37.8) |
| Washed my hands with soap and water more often than usual | 1088/1420 (76.6) |
| Used alcoholic hand gel or hand sanitizer more than usual | 806/1420 (56.8) |
| **Avoidance-related behaviors: Actions taken over last month due to COVID-19 (Cronbach's alpha=0.861)** |  |
| Deliberately cancelled or postponed a social event | 519/1161* (44.7) |
| Cancelled or delayed travelling overseas | 407/811* (50.2) |
| Reduced my use of public transport | 448/880* (50.9) |
| Kept one or more of my children out of school or childcare | 155/587<sup>#</sup> (26.4) |
| Kept away from crowded places generally | 889/1332* (66.7) |
| Increased the amount of household groceries purchased | 453/1377* (32.9) |
| Performed $\geq 1$ of three recommended behaviors | 1205/1420 (84.9) |
| Performed $\geq 1$ of six avoidance behaviors | 1326/1420 (93.4) |

\*Total number of participants lower than overall total of 1420 due to exclusion of participants who ticked 'not applicable' options. <sup>#</sup>587 participants had children of school or childcare going age.

## RESULTS

Of the 1420 respondents, 740 (52%), were female, 47 (3·3%) identified as Aboriginal and/or Torres Strait Islanders, 830 (58·5%) had private health insurance, 792 (55.8%) had children with 211/792 (26·7%) attended childcare/school. The demographic characteristics of the respondents are presented in Table 1. Of the respondents, 37 (%) reported knowing of a COVID-19 case amongst their family or friends. Television news was the primary source of information about COVID-19 (n=724, 51%), followed by government websites (n=241,17%) and social media (n=198, 14%). When asked about the level of trust they had in the information coming from the Government, 667 (47%) stated high to very high.

### Perceptions about susceptibility and severity

Respondents ranked their risk of acquiring COVID-19 as very high (n=71, 5%); high (n=198, 14%), intermediate (n=497, 35%), low (n=397, 28%) and very low (n=156, 11%). The remaining 100 respondents reported not knowing what their risk was. When it came to perceived impact on their health, 710 (50%) reported that COVID-19 would ‘somewhat’ affect it, while the remaining respondents reported it as: extremely (n=170, 12%), seriously (n=326, 23%), not at all (n=85, 6%) or don’t know (n=113, 8%). Fifty percent reported changing their personal perception of risk after reading or hearing information in the media or on social media.

### Perceived effectiveness of social mitigation strategies and ability to adopt

Figure 1 shows the perceptions towards the degree of effectiveness of measures to reduce personal risk from COVID-19. Self-quarantine of anyone who has travelled into Australia from overseas was considered to have high to very high effectiveness (n=1171, 82·5%), followed by avoiding people who have travelled overseas (n=1155, 81·4%). Whereas, only 525 (37%) thought that shutting the restaurants/bars after 6pm would have a high/very high effect and 696 (49%) stated that wearing a mask (when not symptomatic) would be effective. Taking antibiof to very low effectiveness by most (n=908, 64%). Beyond perceived effectiveness, respondents were asked to comment on their ability (self-efficacy) to carry out social distancing strategies, 596 (42%) respondents rated their ability to work from home as high/very high.

**Figure 1:**
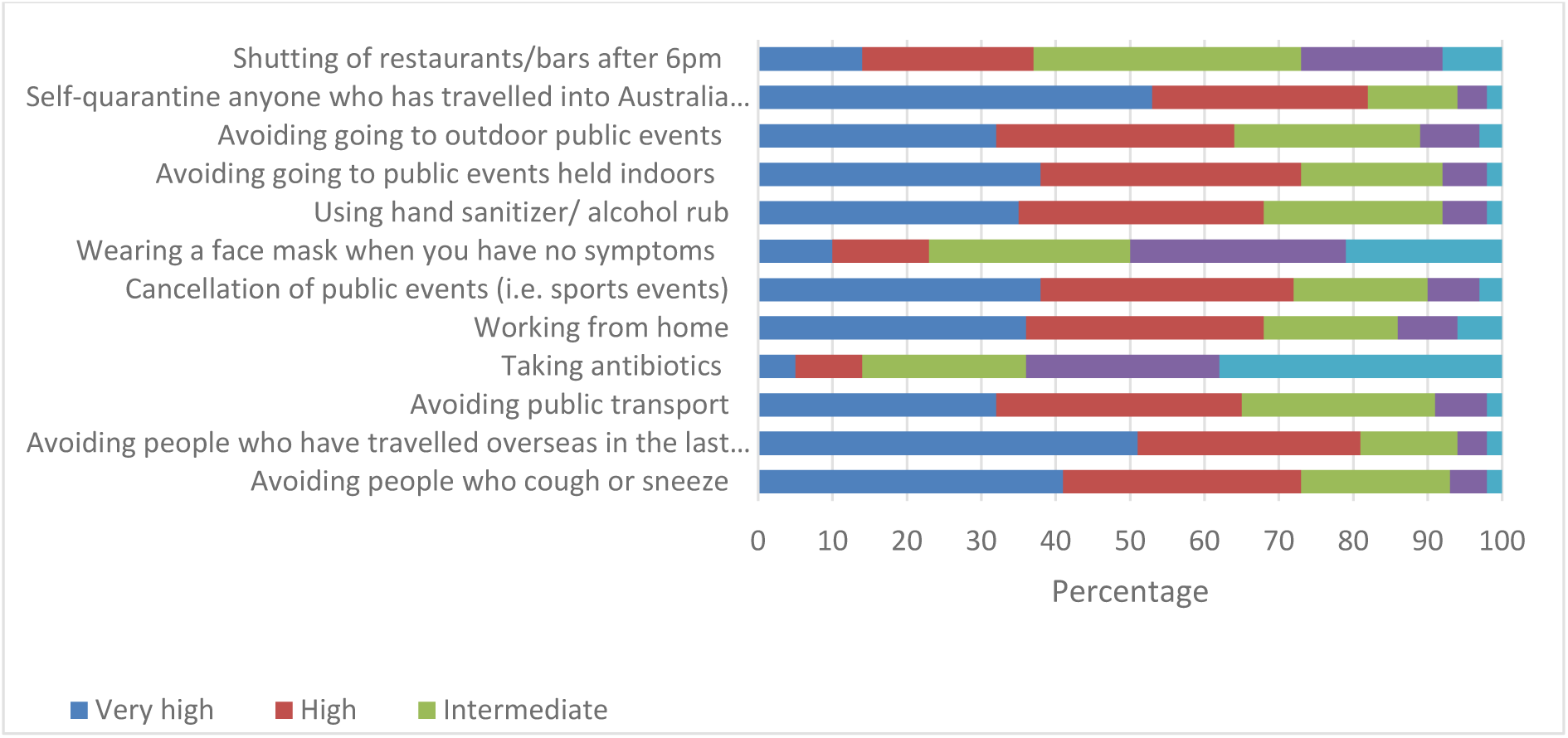
Rating of level of effectiveness of strategies to control Covid-19 outbreak.

### Practice of recommended measures/behavior

The most common hygiene-related behavior adopted was washing hands with soap and water (n=1087, 76·6%), whereas keeping away from crowded places generally was the most common avoidance behavior (n=947, 66·7%). Overall, 1205 (84·9%) respondents reported undertaking ≥ 1 of three hygiene-related behaviors and 1326 (93·4%) performed ≥1 of six avoidance-related behaviors (Table 1). Table 2 shows association between demographic characteristics and reported behaviors during COVID-19 pandemic. Five hundred and five (36·3%) respondents considered the hygiene-related and avoidance-related behaviors as ‘the right thing to do’ as their main motivation to comply. Close to 80% of respondents (n=1127) who reported being worried about COVID-19 (high-very high) were found to have higher engagement with hygiene-related behaviors (OR 3·8, 95% CI: 2·8-5·2) and avoidance-related behaviors (OR 3·1, 95% CI: 2·0-4·8).

**Table 2:**
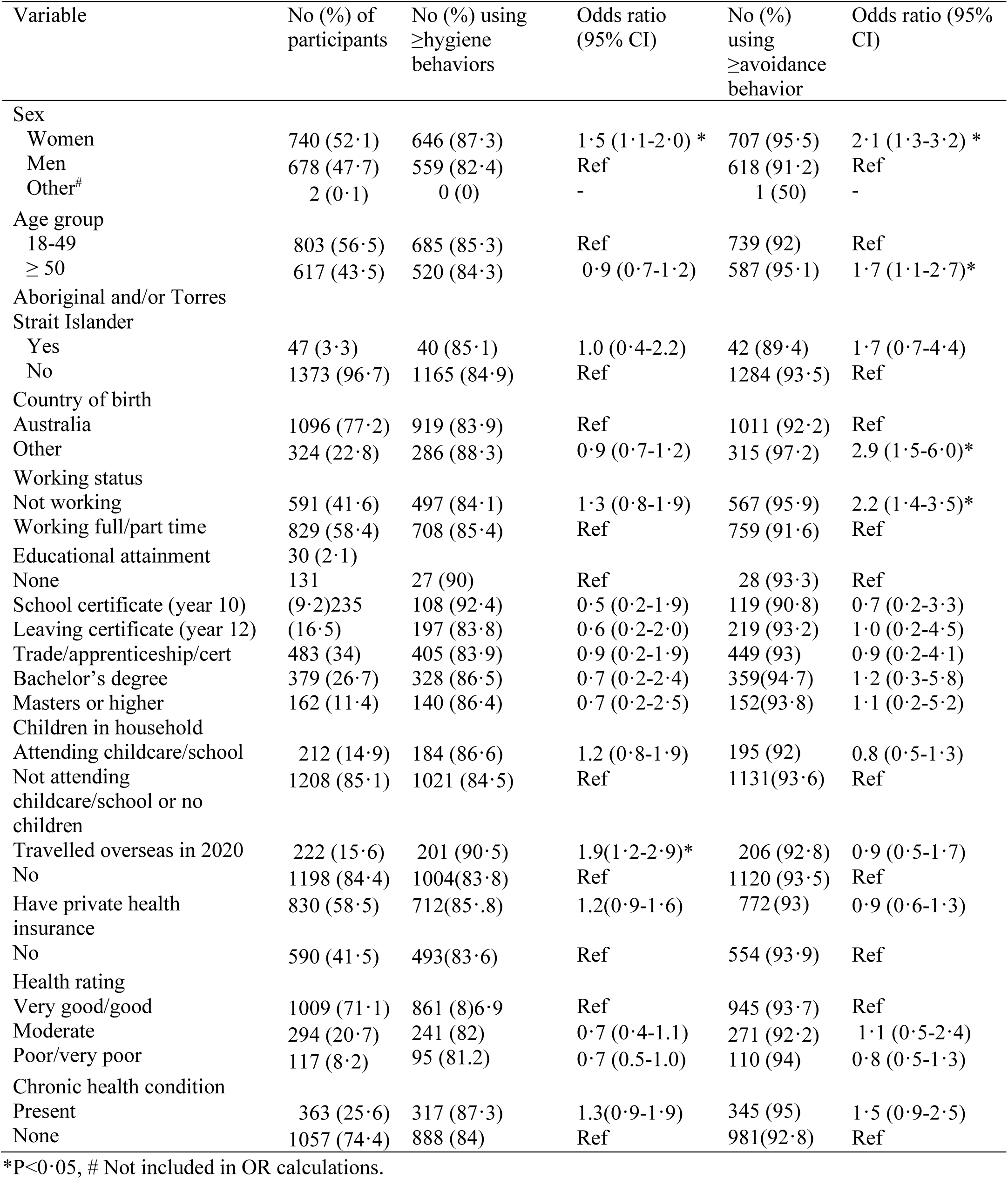
Association between demographic characteristics and adoption of preventive/avoidance strategies during COVID-19 pandemic.

Controlling for demographic and worry variables, there was a higher association between performance of hygiene-related behaviors and trust in government/authorities (aOR: 2·6, 95% CI 1·6-4·4), perceived high severity if infected (aOR: 1·8, 95% CI 1·2-2·7), higher levels of belief in the effectiveness of behaviors (aOR 3·4, 95% CI: 2·5-4·8), higher ability to adopt social distancing strategies (aOR: 1.8,95% CI 1·1-2·1), low levels of concern if self-isolated (aOR: 2·8 95% CI: 1·6-4·8), intermediate to higher level of risk perception (aOR: 2·1, 95% CI: 1·5-2·9, aOR: 3·1, 95% CI: 1·9-4·9) and serious or extreme perceived impact on health (aOR: 1·7, 95% CI: 1·2-2·4) led to performance of recommended behaviors. Reporting the use of avoidance behaviors was more likely in respondents who: trusted government/authorities (aOR: 5·5, 95% CI 3-9·0), rated effectiveness of behaviors higher (aOR: 4.3, 95% CI: 2·8-6·9), and indicated a higher ability to adopt social distancing strategies (aOR: 1.8, 95% CI 1·1-3·0) (Table 3)

**Table 3:**
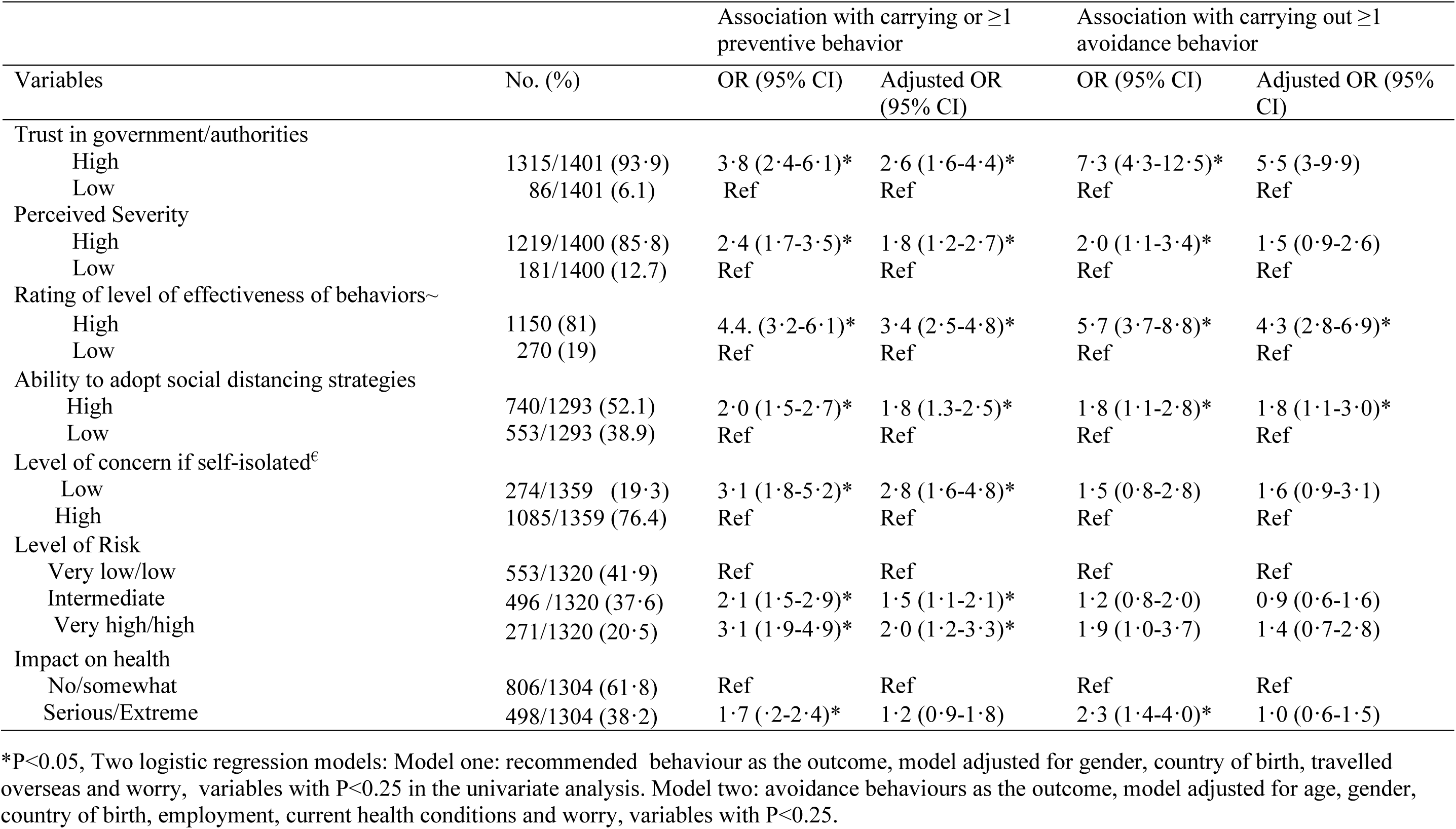
Logistic regression models testing association between perception variables and adoption of hygiene/avoidance strategies during COVID-19 outbreak.

Six questions focused on self-isolation as a strategy. The majority (n=1349, 95%) agreed that they could self-isolate if necessary and that they had a family member or friend who could assist them in the event of isolation (n=1178, 83%). However, respondents did have concerns (high/very high) about not being able to access shops for food/supplies (n=681, 48%) and not being able to access a primary care provider (n=553, 39%). Amongst those who felt they could not manage self-isolation at home (n=122, 8%), the main concerns were centered around carers responsibilities for children, elderly parents and disabled family members.

## DISCUSSION

Our results suggest that a large proportion of respondents have adopted one or more of either the hygiene-related and/or avoidance-related behaviors that had been recommended by the Australian Government. Considering the intense media coverage and government information, it is not surprising that there was a gravitation towards actual (or willingness for) adoption of hygiene strategies including hand washing/sanitizing. While anxiety levels were moderate, concerns were raised about accessing food and medical supplies if placed into self-isolation. Given the limited community behavioral COVID-19 studies published, we have compared our findings to two ongoing community surveys. Firstly, results from an online poll of 14,000 respondents from 14 countries (conducted at the same time point as our study), reported that their respondents in 8/14 countries expressed a belief that social distancing measures such as travel bans, and self-isolation would not prevent the spread of the virus, including participants from Australia (52%) [10]. Secondly, a German survey (conducted a week earlier then our study) identified that respondents had high levels of knowledge, but adoption of important protection behaviors was very low, and risk perceptions were especially low among the elderly [11].

Amongst our participants, perceived susceptibility to COVID-19 was at an intermediate level. There may be several factors at play here that account for the perceived level of risk. Firstly, people may be unaware of true risk since little reporting has focused on attack rates during the time of the study. Secondly, people may be subject to optimism bias – a phenomenon where people downplay their own risk of an outcome[12]. Thirdly, it is likely that some people assess their risk as being low due to already factoring in a change towards anticipated or already accomplished protective behavior [13]. During events that people deem ‘familiar’, we often see unrealistic optimism because the risk is perceived to be under control, as was the case in 2009 with the influenza pandemic, when adoption of precautions was low and there was a sense of personal security [2]. However, COVID-19 presents as an unfamiliar risk (for the large majority of the population had not experienced outbreaks of SARS or MERS) making the risk less tolerable for those who perceive the situation as uncontrollable [14]. When it comes to perceptions of risk, there are numerous studies documenting how they are associated with the uptake of preventive and/or avoidant behaviors. Studies conducted during/after the 2003 SARS outbreak reported that higher levels of perceived risk/susceptibility of SARS was associated with the adoption of preventive behaviors and also avoidance behaviors [15-18].

While understanding a person’s perception of risk is important, it is not the only condition needed for engagement. Higher risk perceptions may only predict protective behavior when people believe that effective protective actions are available (response efficacy) and when they are confident that they can engage in such protective actions (self-efficacy) [19]. According to Bandura social cognitive theory, an individual’s self-efficacy plays a crucial role on the individual’s likelihood to engage in a desired behavior. If an individual does not believe that he/she can carry out the behavior (i.e. physically distance themselves), there is little motivation to engage [20]. Three-quarters of the study respondents agreed that they could adopt the avoidance-related strategies, with lower scores for working from home and self-isolation at home. When asked whether they had adopted any of the hygiene related strategies, washing or sanitizing hands were the most common responses. These represent more readily adoptable strategies, as people in the community understand how to engage in them, believe that the strategy will protect them, and usually have the resources to carry them out. These easy to adopt actions have also been a focus of government mass media messages.

When it came to avoidance behaviors, our respondents were less inclined to rate them as being effective or to have adopted them, in comparison to the preventive behaviors listed above. Perceptions regarding the efficacy of the strategy (as opposed to self-efficacy) have also been found to impact on intentions/likelihood to adopt or actual uptake [17, 21]. It is not surprising that some strategies including social distancing, scored low as people may not understand what the strategy entails, the rationale for its use, or what impact it may have on one’s health. It should also be noted that individuals may not have the capacity or resources to comply with physical distancing measures because they: (1) have extended families living in their households; (2) they have a responsibility to provide care for someone outside of their home; (3) they may reside in share accommodation; (4) may not have access to internet/computer in the home setting or (5) because of the type of job they have they cannot simply shift to working from home. In these settings, providing mass media education is not going to suffice. What is needed is pragmatic solutions that support people financially and socially to participate. Examples including increases in social support and charities to assist with delivery of groceries and meals, home delivery from chemists, telehealth consultations bulk billed, drive through vaccine clinics etc.

It has been suggested that we start to prime people about what additional strategies may still need to be introduced [22]. This would entail talking with them about why the strategy would be implemented; the end-goal of implementing it; what could be the potential impacts; how members of the public engage; and the criteria for its de-escalation. In order to promote cooperation with social (physical distancing) strategies, governments may need to use realistic portrayals (community stories) and role modelling by influential actors in social networks. Observing competent role models perform actions that result in success conveys information to observers about the sequence of actions to use to be successful [23]. Motivation may be helped by creating media campaigns that foster awareness of the recommended behaviors and encourage people to share their strategies for complying with self-isolation and working from home.

Amongst our respondents, older age was associated with the adoption of precautionary behaviors, which aligns with the findings from Singapore and Hong Kong during the 2003 SARS outbreak [5, 17] and some studies during the 2009 H1N1/A pandemic [24, 25]. However, the pattern of age is not straight forward. In contrast to the above studies, others have reported higher levels of adoption of preventive behaviors amongst younger people (18-24 years) in the context of the 2009 influenza pandemic. When it comes to gender, we found that females were more likely to report uptake of both preventive and avoidance behaviors, consistent with studies during SARS and H1N1 pandemic influenza [2, 5, 17, 26]. Earlier studies have indicated that women are more likely to perceive themselves to be susceptible and hence adopt the behaviors [15, 27]. When it comes to country of birth, we found that people born outside of Australia were less likely to adopt behaviors. This finding may relate to the capacity to access information, which at the time, was being disseminated in English and largely through mainstream media conferences and health department websites rather than community and language groups. Further work is needed to explore the associations between country of birth and pandemic-related behaviors.

When asked what would motivate respondents to comply with a social distancing strategy, they nominated ‘I believe it is the right thing to do’ as the primary response. While this answer did not have any significant relationship with the outcome measures reviewed, it is still relevant when it comes to planning communication messages. It suggests that respondents may be influenced by a desire for social approval from others, an idea linked to the model of moral motivation. The model, developed by Brekke et al. (2003), assumes that individuals have preferences for achieving and maintaining a self-image as a socially responsible person [28]. In Brekke et al.’s model, self-image improves when the individual’s actual behavior gets closer to her/his view of the “morally ideal” behavior (i.e. the behavior that would maximize social welfare if chosen by every member of society). However, individual’s participation can be conditional on whether they think others are also contributing [29]. Mass media campaigns that frame their messages around a social collective action/power or the inclusion of the general public within a team to assist the community response may be effective. The promotion of prosocial behaviors has been shown to be effective in vaccination uptake and could be adapted in promoting COVID-19 mitigation behaviors, such as how one’s actions can contribute to protecting their grandparents [30]. This idea has been picked up by celebrities and the wider community on twitter under the #LockDownForLove, with people nominating who they are social distancing for. Whether these strategies work, needs to be further examined.

Early results from a study conducted across the US, UK and Germany has suggested that inducing empathy for those most vulnerable to the virus promotes the motivation to adhere to physical distancing [31]. The use of empathy in messaging is not a new concept and has been applied in a range of ways from the promotion of testing/treatment for STIs, through to increasing our acceptance of robots [32, 33]. Empathy is a skill which enables understanding of another person’s experience. Here, people could be asked to imagine the perceptions, needs and impact (health, financial, social) of pandemic COVID-19 amongst our family members/friends.

Our study includes a large, representative cross-section of the adult Australian population. People who could not communicate in English were excluded from the sample, which may have affected representation of ethnic minorities. We also had under-representation of Aboriginal Australians and Torres Strait Islanders and from those in remote settings. Secondly, as participation in our study was on a voluntary basis, this study has potential for self-selection bias by community members who are particularly concerned about this pandemic. In addition, we relied on self-reports of behaviors which may have led to over-reported (social desirability bias). However, this may have minimized as the survey was self-complete and anonymous.

Based on the available data, it appears that older individuals (aged >60 years) and people with chronic underlying health conditions are particularly susceptible to severe disease. This presents a challenging situation. In the media there is reporting that ‘COVID-19 is causing mild illness’ in the majority but it’s in the best interest of the country to stay home in order to ‘flatten the curve’. This will cause two responses – those who continue with their normal practice (not adopting or complying with the recommendations around social distancing/mitigation strategies) as identified in a proportion of our respondents. Motivating this group (especially those less likely to be at risk or suffer the health impact of COVID-19) to adopt behaviors that require marked change in their routines, beyond those related to personal hygiene. It may prove difficult unless people understand the required behavior, the rationale for it, are given clear and sufficient information about how to comply, and they believe the strategy will have an impact and are motivated to act [7, 17, 34]. They also need to have capacity and opportunity to comply with new behaviors for another 4 to 6 months. In order to engage a community, they need to feel like they are a valued part of a team, and that their contributions are valued and key to the response. Lastly, it is essential that governments ensure that resources, legalization and support measures are in place in order to facilitate community participation in community mitigation strategies.

## Data Availability

The data is available upon request

## DECLARATIONS

### Authors’ contributions

HS conceived the study, undertook the data collection and developed the journal paper. All other authors assisted with the design, interpretation and with revising the paper. All authors have read and approved the manuscript.

## Acknowledgments

We would like to thank the respondents for their time in participating in the research study. MS is supported by a fellowship from the Westpac Scholars Trust.

## Supplementary materials

**List of original and recoded predictor variables**.

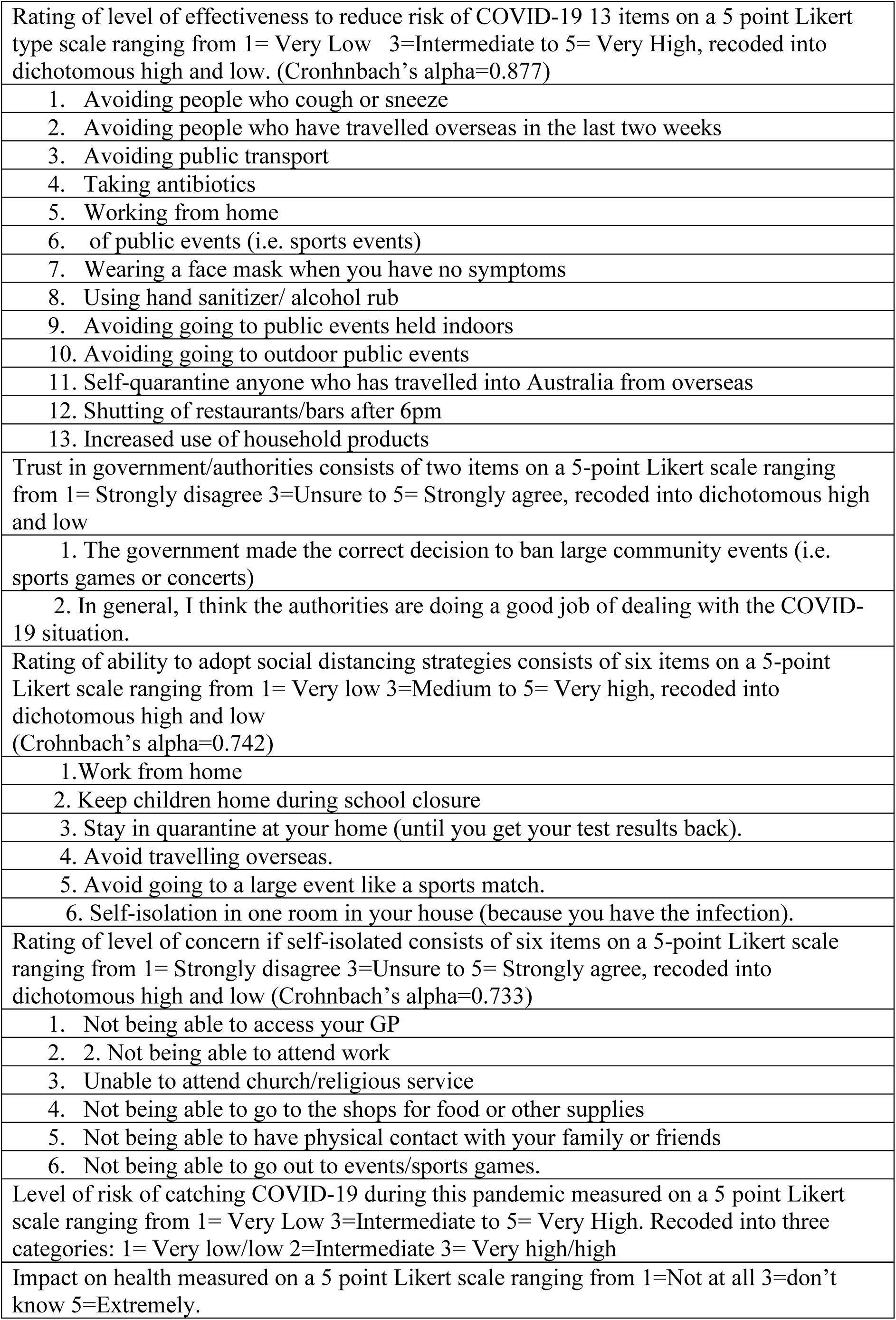

